# Comprehensive Exome Sequencing in Swedish Patients with Spontaneous Coronary Artery Dissection

**DOI:** 10.64898/2026.04.22.26351535

**Authors:** Cecilia Gunnarsson, Rada Ellegård, Josefine Åhsberg, Sophia Huda, Jonas Andersson, Christian Dworeck, Natalie Glaser, David Erlinge, Loghman Henareh, Nina Johnston, Maria Mannila, Christos Pagonis, Annica Ravn-Fischer, Erik Rydberg, Kerstin Welén Schef, Per Tornvall, Eva Swahn, Sofia Sederholm Lawesson

## Abstract

**Background:** Spontaneous coronary artery dissection (SCAD) is a well-recognised cause of acute coronary syndrome particularly among women without conventional cardiovascular risk factors. Increasing evidence indicates a genetic contribution; however, the underlying genetic architecture of SCAD remains insufficiently understood.

**Objective:** The aim of this study was to assess the prevalence of rare variants in previously reported SCAD associated genes and to explore the potential presence of novel genetic alterations in well-characterised Swedish patients with SCAD.

**Methods:** The study comprised 201 patients enrolled in SweSCAD, a national project examining the clinical characteristics, aetiology, and outcomes of SCAD. All individuals had a confirmed diagnosis based on invasive coronary angiography. Comprehensive exome sequencing was performed to identify rare variants contributing to disease susceptibility.

**Results:** Genetic variants that have been associated with SCAD according to current clinical genetics practice for variant reporting were identified in approximately 4 % of patients. In addition, rare potentially relevant variants were detected in almost 60 % of patients in genes associated with vascular integrity and vascular remodelling.

**Conclusion:** This study supports SCAD as a genetically complex arteriopathy, driven by rare high-impact variants together with broader polygenic susceptibility. Variants in collagen, vascular extracellular matrix, and oestrogen-responsive pathways provide biologically plausible links to female-predominant disease. Although the diagnostic yield of clearly actionable variants is modest, these findings support broader genomic evaluation beyond overt syndromic presentations and highlight the need for larger integrative genomic and functional studies to refine risk stratification and management.

## Introduction

Spontaneous coronary artery dissection (SCAD) is a non-atherosclerotic, non-traumatic cause of acute coronary syndrome that predominantly affects middle-aged women, many of whom lack conventional cardiovascular risk factors^1^. Although SCAD accounts for a minority of all myocardial infarction (MI) cases, it comprises over 30% of the cases in women under 60 ^2^.The pathogenesis is thought to involve an underlying arteriopathy that results in increased fragility of the coronary arterial wall. In susceptible individuals, a precipitating trigger may provoke a dissection, arising either from an intimal tear or from spontaneous intramural haemorrhage, typically localised within the tunica media. Subsequent compression of the true lumen leads to myocardial ischaemia and ultimately MI^1^.

Although the underlying mechanisms remain incompletely understood, both environmental and genetic factors are believed to contribute to disease susceptibility, including hormonal fluctuations, physical exertion, and emotional stress. Previous studies have shown that patients with SCAD frequently exhibit concomitant vasculopathies, including fibromuscular dysplasia (FMD), extracoronary dissections, aneurysms, and migraine^3^.

A potential genetic predisposition to SCAD has been highlighted in earlier studies, with several rare and common variants identified in genes implicated in connective tissue organisation, extracellular matrix integrity, and vascular smooth muscle function^4–7^. Sequencing studies focusing on rare variants have demonstrated that many variants are harboured in genes known from hereditary connective tissue disorders (CTDs), such as vascular Ehlers–Danlos syndrome (vEDS), Loeys–Dietz syndrome (LDS), Marfan syndrome, and autosomal dominant polycystic kidney disease. Potentially pathogenic variants in genes associated with vasculopathies or CTDs have been identified in approximately 3–10 % of individuals with SCAD^7,8^.

The objective of this study was to assess the prevalence of rare variants in previously characterised SCAD associated genes and to explore the potential presence of novel genetic variants associated with vasculopathy in a well-defined Swedish group of patients with SCAD. A deeper understanding of the complex genetic background of SCAD may help identify patient subgroups who could benefit from targeted clinical genetic testing, enabling more targeted follow-up for both patients and their families.

## Material

This study includes Swedish patients with a clinical diagnosis of SCAD included in SweSCAD, a national project designed to investigate diagnostics, aetiology, associated connective tissue and vascular disorders, treatment, prognosis, and long-term well-being following SCAD. The inclusion criterion was a SCAD diagnosis confirmed by invasive coronary angiography (ICA). Patients were enrolled via three routes:

1. **Prospective recruitment** during the index SCAD hospitalisation at seven Swedish hospitals (n=46, of whom 40 consented to participate in the present study).
2. **Retrospective recruitment** at outpatient clinics in eight hospitals (n=118, of whom 76 consented).
3. **Identification through the Swedish Coronary Angiography and Angioplasty Register (SCAAR)** between 2015 and 2017, followed by ICA re-evaluation and confirmation of SCAD (n=147, of whom 85 consented)^9^.

After providing written informed consent, participants received a questionnaire addressing comorbidities, pregnancy/post-partum status, potential trigger events, and medication use. Each participant was also provided with a DNA Genotek saliva collection kit and written instructions for self-sampling to be performed within one year of enrolment. Saliva samples and signed consent forms were returned to the coordinating centre in Linköping using prepaid packaging. All samples were assigned a unique study identifier. The study was approved by Swedish Ethical Review Authority (#2020-00748) according to the declaration of Helsinki. All participants provided written informed consent.

## Methods

DNA extraction was performed using the Maxwell® RSC Stabilized Saliva DNA Kit according to the manufacturer’s instructions. Exome libraries were prepared using the Twist Human Core Exome plus RefSeq panel and sequenced on an Illumina NextSeq 500 platform following standard protocols. Sequence alignment and variant calling were conducted using DNAseq (Sentieon) with GRCh37 as the reference genome, and variants were annotated using Emedgene (Illumina). Copy number variant (CNV) detection was performed using ExomeDepth^10^.

Variants were filtered using three complementary strategies:

- **AI-based ranking:** Emedgene’s AI algorithm was queried using the HPO term HP:0006702 – Coronary Artery Dissection, and all candidate variants flagged by the algorithm were evaluated.
- **Literature gene panel:** All variants identified in genes previously reported to be associated with SCAD were reviewed based on a literature-derived gene list.
- **Manual filtering:** Variants with low population frequency and predicted deleterious effect by *in silico* tools were assessed.

A schematic overview of the analysis strategy is provided in Figure 1.

**Figure 1.**
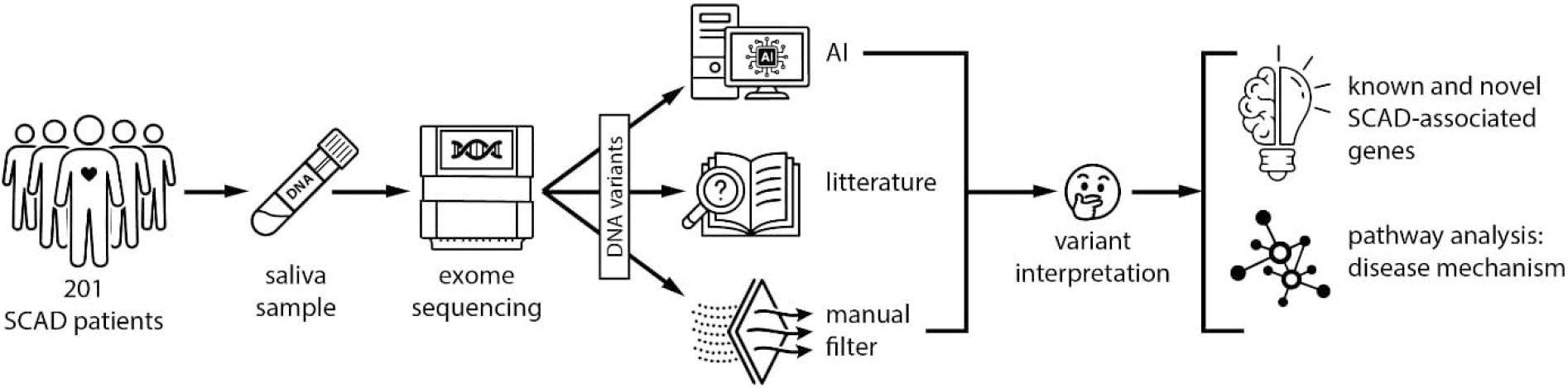
Study overview. Exome sequencing was performed on samples from 201 SCAD patients. The genetic variants were filtered using three different strategies: ranking with an AI supplied with the HPO term “coronary artery dissection”, filtering on a gene panel including genes previously reported to be associated with SCAD, and a manual filter based on variant population frequency and variant effect prediction algorithms. The resulting variants were manually reviewed and classified as “clinically reportable”, “strong”, “medium”, “weak” candidate variants or as non-relevant. Variants scored as “medium” or stronger were found in both previously reported and novel genes. These genes were subjected to pathway analysis to gain insight into SCAD disease mechanisms.

The literature search consisted of a review of all publications indexed in Web of Science using the query “SCAD genetics”. Variants with a population frequency <0.05 in gnomAD v4.1.0 and in an internal historical database (~1500 clinical exomes processed using the same library chemistry) were included for evaluation.

For manual filtering, rare variants were defined as those with a prevalence <0.05 in both gnomAD v4.1.0 and the internal database. Potentially deleterious loss-of-function variants were defined as truncating or splice-affecting variants (SpliceAI score >0.2) in genes intolerant to loss of function (gnomAD pLI >0.9 or LOEUF <0.6)^11^. Potentially deleterious missense variants were defined as those with a REVEL score ≥0.644^12^.

The objectives of the variant filtering were twofold: first, to identify clinically reportable and disease-causing variants for SCAD according to current clinical practice; and second, to detect novel genetic findings without an established link to vasculopathy. All variants identified through the filtering strategies were manually reviewed, interpreted, and classified as “clinically reportable”, “strong”, “medium”, or “weak” candidate variants, or dismissed as non-relevant.

“Clinically reportable” variants were defined as those occurring in genes with a recognised association with SCAD that would be reported in routine clinical diagnostics under current practice. Interpretation followed the consensus guidelines of the American College of Medical Genetics and Genomics (ACMG) and the Association for Molecular Pathology^13^. A physician specialising in clinical genetic diagnostics (C.G.) determined which variants qualified as clinically reportable. Patients with clinically reportable variants were handled according to clinical guidelines.

Variants in genes without an established phenotype association cannot be classified as pathogenic or likely pathogenic according to ACMG criteria. Consequently, all candidate variants not deemed clinically reportable were categorised as variants of uncertain significance (VUS). These were further subclassified into “strong”, “medium”, or “weak” variants, or dismissed, based on an integrated assessment. This evaluation included allele frequency in gnomAD, gene constraint metrics, the number and nature of variants reported in ClinVar, and available functional information.

Although coronary artery gene expression (GTEx) was reviewed, this parameter was given limited weight, as many genes previously implicated in SCAD demonstrated low expression levels (15% <2 TPM; 27% <10 TPM in coronary artery tissue). All variant filtering and classification were performed by a single experienced interpreter (R.E.). STRING analysis for GO Cellular Component was performed using ShinyGO 0.85.1^14^. Pathway analysis was conducted using IPA (Qiagen), complemented by an extensive literature review.

## Results

A total of 201 patients were included in the study; patient demographics are shown in Table 1. In total, 5,582,339 SNP/indel variants were detected across the patient group (see Supplementary Table 1 for all identified candidate variants). For downstream analyses, only candidate variants classified as medium or stronger were retained.

**Table 1.**
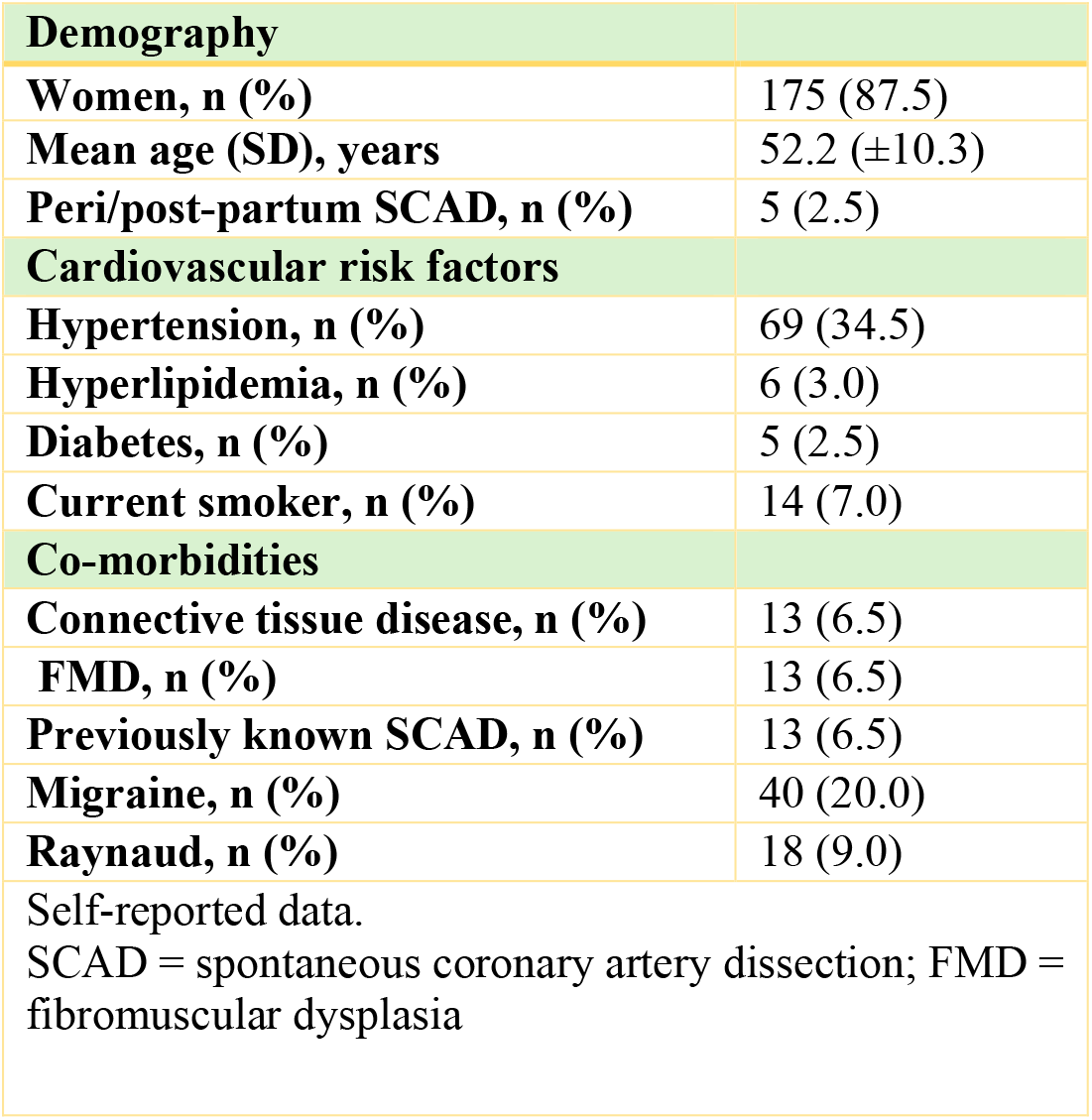
Basic characteristics.

| **Demography** |  |
| --- | --- |
| **Women, n (%)** | 175 (87.5) |
| **Mean age (SD), years** | 52.2 (±10.3) |
| **Peri/post-partum SCAD, n (%)** | 5 (2.5) |
| **Cardiovascular risk factors** |  |
| **Hypertension, n (%)** | 69 (34.5) |
| **Hyperlipidemia, n (%)** | 6 (3.0) |
| **Diabetes, n (%)** | 5 (2.5) |
| **Current smoker, n (%)** | 14 (7.0) |
| **Co-morbidities** |  |
| **Connective tissue disease, n (%)** | 13 (6.5) |
| **FMD, n (%)** | 13 (6.5) |
| **Previously known SCAD, n (%)** | 13 (6.5) |
| **Migraine, n (%)** | 40 (20.0) |
| **Raynaud, n (%)** | 18 (9.0) |
| Self-reported data.<br>SCAD = spontaneous coronary artery dissection; FMD = fibromuscular dysplasia |  |

### Identification of candidate variants

AI-based screening identified 389 variants in 129 genes, of which 7 were classified as clinically reportable, 5 as strong, and 4 as medium candidate variants, the remaining 373 variants were judged to be weak or non-relevant. The literature-based strategy identified 128 genes with previously reported associations with SCAD (Supplementary Table 2). Among these genes, 756 rare variants were detected and evaluated. This resulted in 8 clinically reportable, 33 strong, and 30 medium candidate variants. Candidate variants of medium and strong strength were found in 46 of the 128 previously implicated SCAD-associated genes.

The manual filtering strategy screened all genes for variants with a population frequency <0.05 and predicted deleterious effect based on variant effect prediction algorithms. 2453 variants were identified: of these, 8 were classified as clinically reportable, 83 as strong, and 68 as medium candidate variants.

All but one of the clinically reportable variants were detected by all three strategies. The clinically reportable variants were all unique, and no patient had more than one clinically reportable variant. Clinically reportable variants were found in *COL3A1* (2 females), *FBN1* (1 female, 1 male), *FBN2* (1 female), and *SMAD3* (2 females, 1 male). According to ACMG criteria, 4 variants were classified as Pathogenic/Likely Pathogenic and 4 were classified as variants of unknown significance (VUS). The *FBN2* variant was identified through both the literature-based and manual filters but did not appear on the AI-generated candidate list. All clinically reportable variants and their ACMG classifications are listed in Table 2.

**Table 2.** Clinically reportable variants. Identified rare variants that would have been reported clinically according to current clinical routine using ACMG criteria (American College of Medical Genetics and Genomics)^14^. (MANE= Matched Annotation from NCBI and EBl, VUS = Variant of Unknown Significance)

| Gene | c. pos (MANE) | p.pos | ACMG criteria | ACMG classification |
| --- | --- | --- | --- | --- |
| *COL3A1* | c.2356G>A | p.Gly786Arg | PM2_Mode+PP3_Strong+PM5_Moderate | Pathogenic |
| *COL3A1* | c.2587C>T | p.Arg863Cys | PM2_Moderate+PP3_Moderate | VUS |
| *FBN1* | c.802T>C | p.Ser268Pro | PM2_Supporting+PM1_Moderate+PP3_Supporting+PP2_Supporting | VUS |
| *FBN1* | c.5423-2A>G | p.? | PM2_Supporting+PVS1_VeryStrong | Likely Pathogenic |
| *FBN2* | c.1525C>T | p.Arg509Cys | PM2_Supporting + PP3_Supporting | VUS |
| *SMAD3* | c.221G>A | p.Arg74Gln | PM2_Moderate+PM5_Moderate+PP3_Moderate | Likely Pathogenic |
| *SMAD3* | c.238C>T | p.Arg80Trp | PM2_Moderate+PP3_Moderate | VUS |
| *SMAD3* | c.1178C>T | p.Pro393Leu | PM2_Moderate+PP3_Strong | Likely Pathogenic |

Each of the three strategies generated unique strong and medium candidate variants not identified by the other approaches (Figure 2; Supplementary Table 1).

**Figure 2.**
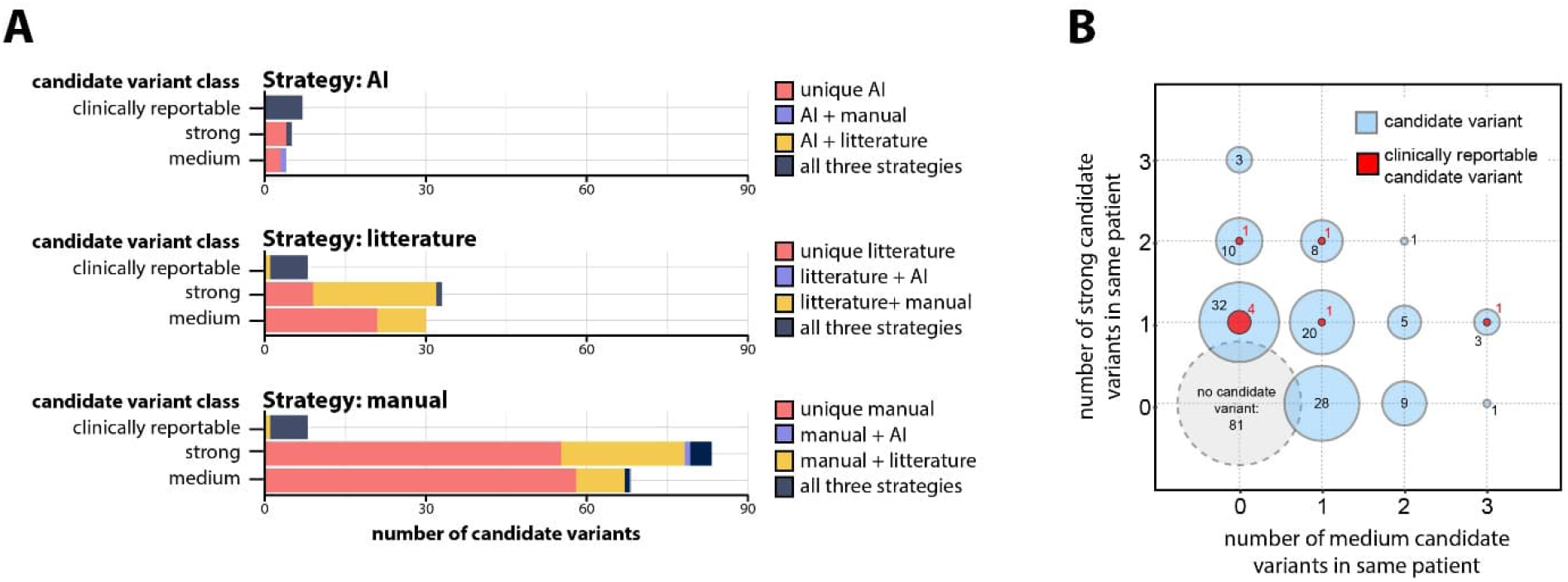
Number of identified candidate variants. **(A)** Number of candidate variants classified as at least strength “medium” identified through the three candidate variant identification strategies. **(B)** Number of candidate variants co-occurring in the same patient.

Copy-number variants (CNVs) were analysed separately, and one CNV-COL4A1 exon 16 deletion was classified as a strong candidate variant.

A candidate variant of strong strength was identified in 83 patients (41.3% of the cohort, 72 females, 11 male), including 8 individuals harbouring clinically reportable variants (4.0%, 6 female, 2 male). A candidate variant of at least medium strength was identified in 120 of the 201 patients (58.7%, 15 male, 105 female). Sixty patients carried a single candidate variant, and the same number harboured two or more candidate variants (Figure 2B).

### Pathway analysis

STRING analysis revealed enrichment of variants in genes associated with Collagen-containing extracellular matrix (41 variants in 21 genes), Focal adhesion (27 variants in 18 genes), Sarcomere (25 variants in 13 genes), Actin cytoskeleton (28 variants 18 genes) and Cation channel complex (12 variants 8 genes). For the complete STRING analysis, see Supplementary Table 4.

All genes with variants of at least medium strength were uploaded to Ingenuity Pathway Analysis (IPA) to identify common upstream regulators potentially central to SCAD pathogenesis. The five most significant upstream regulators were TGFB, angiotensinogen, beta estradiol, liposaccharide and TNF (Figure 3). Upstream regulators are shown in Figure 4, with regulated genes listed in Supplementary Table 3. All genes containing a candidate variant of at least medium strength were also linked into a unified pathway using IPA complemented by an extended literature review (Figure 4). No significant differences in genetic findings related to specific signalling pathways were observed in association with clinical characteristics such as migraine, sex, or fibromuscular dysplasia (FMD).

**Figure 3.**
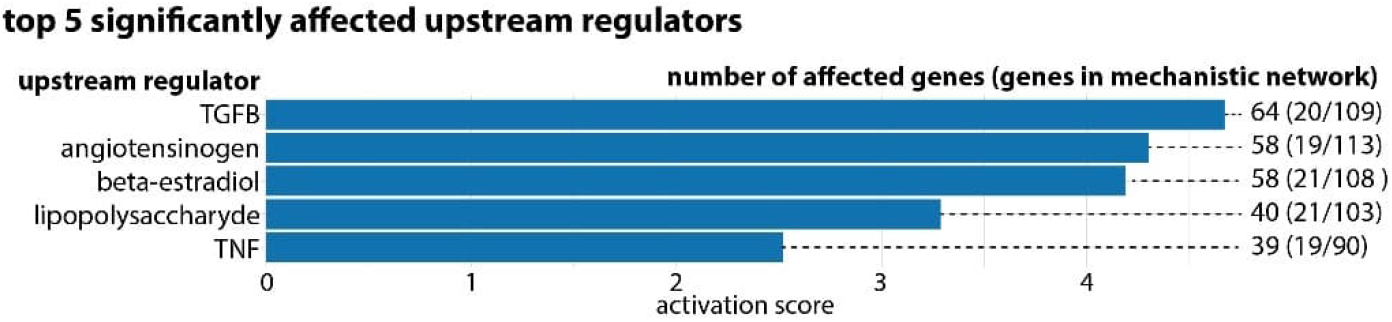
Top upstream regulators of candidate genes. Genes harboring a candidate variant of at least strength “medium” were subjected to pathway analysis (IPA) to identify which upstream regulators were associated with the affected genes. Numbers denote number of genes known to be affected by regulator, numbers in brackets denote number of these genes in IPAs mechanistic network (of total in the network). The same gene can be counted as affected by several upstream regulators and can be part of several mechanistic networks.

**Figure 4:**
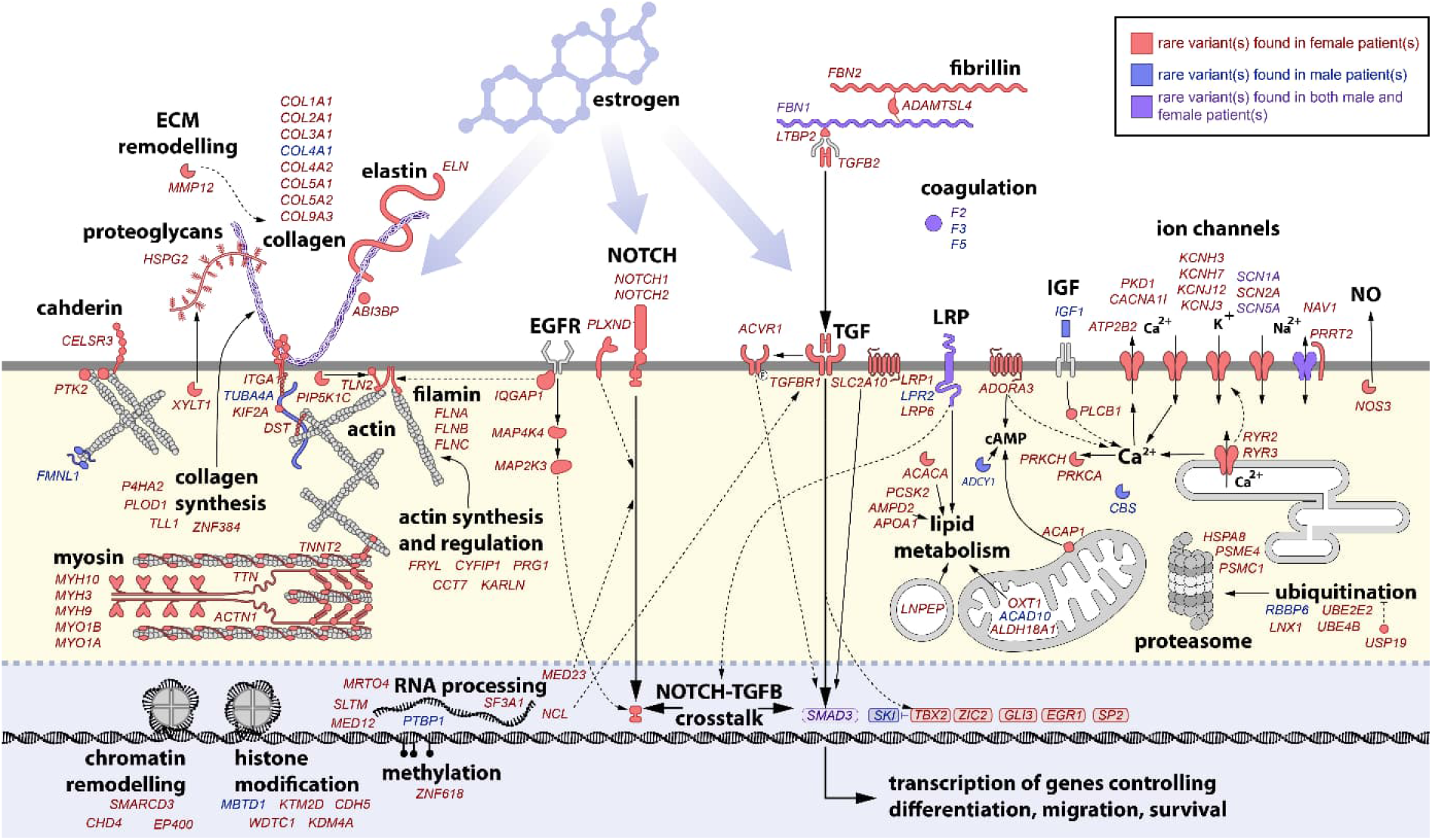
Genetic variants classified as ‘clinically reportable’, strong’ or ‘medium’ were connected in a pathway using IPA and literature studies, presented to illustrate which biological processes are considered to be affected among SCAD patients in the present study.

## Discussion

In this study, we performed exome sequencing on 201 Swedish patients with SCAD and identified clinically reportable variants in approximately 4% of cases, a finding consistent with prior reports. Patients carrying rare pathogenic variants associated with aortic or connective tissue disorders sometimes do not present with the classical clinical features typically observed in conditions such as vEDS, LDS, or Marfan syndrome. The absence of a clear phenotypic profile suggests that restricting genetic testing to patients exhibiting overt connective tissue manifestations may result in missed diagnoses. This observation supports the consideration of broader genetic evaluation in SCAD, despite the current diagnostic yield using clinically available gene panels remaining between 3% and 10%^1,2^. Although consensus guidelines for routine genetic testing in SCAD have not yet been established, genetic testing may be considered appropriate for selected patient groups as the results could have high impact on the specific individual and their family even if the diagnostic yield on group level is low.

To elucidate the genetic basis of SCAD, several approaches have been employed. Genome wide association studies (GWAS) have examined the contribution of common genetic variants^5^, whereas whole exome and whole genome sequencing (WES/WGS) studies have focused on rarer genetic determinants^7,8^. As rare variants associated with a monogenic disease mechanism are typically found in only a small subset of SCAD patients, a model of complex genetic inheritance has been proposed^15,16^. In other disorders, digenic inheritance models have been implicated in disease pathogenesis, and similar mechanisms may also contribute to SCAD development^17^. It is plausible that SCAD arises from a spectrum of genetic influences, ranging from high impact, highly penetrant variants absent from the healthy population to additive effects of multiple common variants, each exerting modest individual effects.

In the present study, we chose to focus on rare variants predicted to have high functional impact (Figure 5A), as these variants include those currently reportable in clinical practice and may influence patient management. High impact variants are also valuable for elucidating the core molecular processes underlying SCAD pathology (Figure 5B).

**Figure 5.**
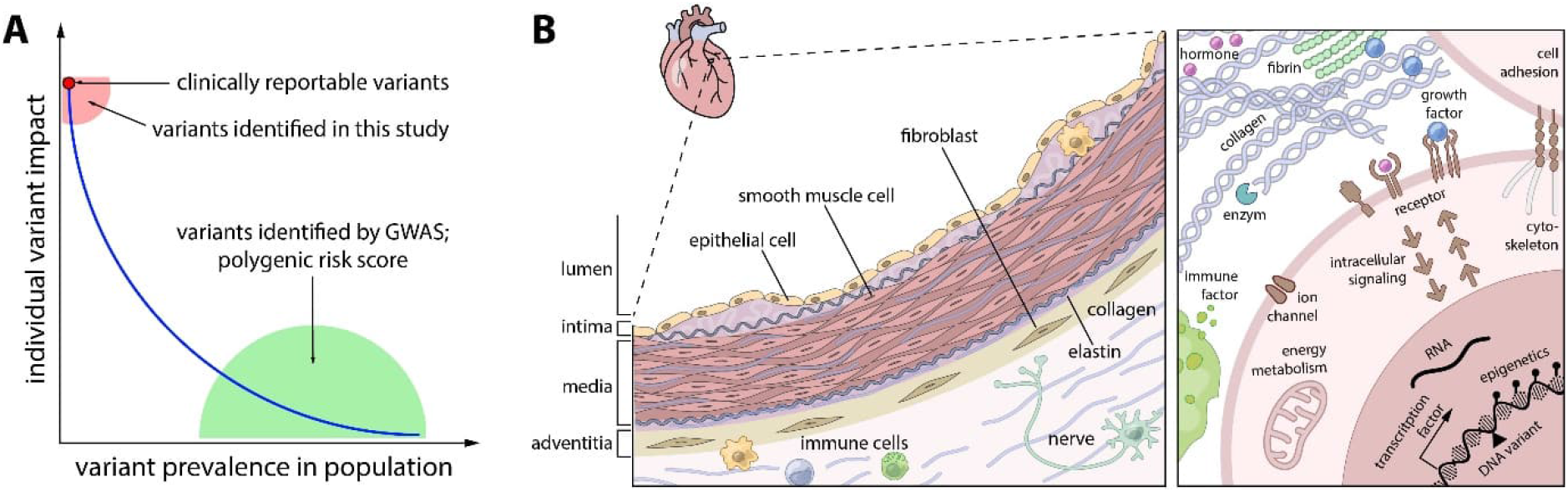
The genetics of SCAD. **(A)** Predisposition to SCAD can be caused by everything from one single genetic variant not present in the healthy population with a high impact, to the additive effect of many different variants common in the population that individually have a low impact. In this study, we have focused on identifying rare variants with high impact. **(B)** Different factors contribute to the stability of the coronary artery wall, and dysregulation leading to SCAD can be associated with defects in various genes. Both intra- and extracellular proteins are involved, as well as factors influencing their expression and regulation.

Women are disproportionately affected by SCAD, accounting for approximately 90% of cases, with increased vulnerability during the peripartum period and menopause, physiological states characterised by reduced oestrogen levels^4,5^. Oestrogen confers protective effects on the vascular endothelium and stromal tissue^18^, and in our study we identified rare variants in 58 genes reported to be modulated by oestrogen signalling. Previous studies have indicated that intact NOTCH signalling is essential for oestrogen-mediated vasoprotection^19,20^. In the present study, we observed genetic variants within genes involved in the NOTCH signalling network. This raises the possibility that congenital susceptibility, such as altered NOTCH signalling, might potentiate vascular injury in the context of fluctuating oestrogen levels, including peripartum or perimenopausal periods. Furthermore, we detected multiple variants in genes implicated in the NOTCH/TGF crosstalk pathway that are not represented in healthy population databases. For example, we identified variants in GLI3, a gene previously demonstrated to regulate angiogenesis and endothelial cell activity^21^. Variants were also found in SKI, and earlier work has shown that SKI overexpression can impair TGF-β signalling in a manner similar to mutations in TGF-β receptors or SMAD proteins, implicating it in connective tissue and endothelial integrity^22,23^.

Zekavat et al. performed WES on 130 SCAD patients and demonstrated a significantly increased burden of variants in collagen-related pathways compared with healthy controls^24^. In our study, we similarly identified rare variants in genes related to collagen biology. However, in both studies, application of ACMG guidelines resulted in many variants being classified as variants of uncertain significance (VUS). This underscores a fundamental challenge in SCAD genetics, distinguishing pathogenic variants from rare benign variation.

Most variants identified here currently lack sufficient evidence to be used in clinical diagnostics, highlighting the need for deeper functional characterisation. These findings support a model in which such variants confer increased susceptibility that, in combination with environmental or hormonal triggers, contributes to disease expression. Using our classification approach, we identified potentially relevant variants affecting vascular integrity in 60% of patients. This supports the concept that a “second hit,” or partial impairment in collagen or other structural pathways, may be required for disease manifestation. A recent study identified variants in COL4A1 and COL4A2 associated with SCAD and related vascular disorders^5,8,25^. These genes encode type IV collagen α-chains, essential components of the vascular basement membrane. Disruption of collagen biosynthesis or structure can compromise extracellular matrix stability, leading to increased vascular fragility^19,26^. Furthermore, Zekavat et al. developed a murine model of partial collagen deficiency, demonstrating mild aortic dilatation and spontaneous arterial dissections, phenotypes more pronounced in females^24^. Transmission electron microscopy revealed aberrant collagen fibril architecture and reduced fibril diameter, emphasising the direct impact of collagen gene variation on vascular integrity. Our findings support a genetically complex model involving both rare monogenic contributors and polygenic or oligogenic mechanisms.

Migraine is highly prevalent in SCAD, with studies reporting prevalence rates between 37 % and 46 %, substantially higher than in the general female population^27,28^. Endothelial dysfunction associated with migraine is thought to contribute to vascular pathologies including stroke and cervical artery dissection, with similar mechanisms proposed for SCAD^28^. Twenty percent of the patients included in the present study reported a diagnosis of migraine; however, no genetic differences were observed between individuals with and without self-reported migraine in our cohort. We observed that a small number of patients with collagen-pathway variants also harboured variants in ion-channel genes, raising the possibility of digenic inheritance, a mechanism in which neither variant alone is sufficient to cause disease, but their combined effect increases susceptibility^17^.

Fibromuscular dysplasia (FMD) is widely recognised as a major predisposing condition for SCAD, and the relationship between these conditions continues to be actively investigated^29–31^. SCAD frequently co-occurs with extracoronary vascular abnormalities, suggesting a systemic arteriopathy^3,32^. In this study of patients with self-reported FMD, we were unable to identify any genetic differences between those with SCAD and FMD and SCAD without FMD. However, the number of participants who reported FMD was very small in the present cohort. Future studies should therefore ensure that the diagnosis of FMD is objectively and clinically confirmed.

### Strengths and Limitations

A key strength in the present study is the national, well-validated cohort with angiography-confirmed SCAD, combined with a robust multi-method genetic analysis, increasing both reliability and clinical relevance, however the study has several limitations. The cohort size was modest and restricted to a Swedish population, potentially limiting generalisability to broader populations. Interpretation of genetic variants remains challenging, with a high proportion classified as variants of uncertain significance, complicating clinical decision making and emphasising the need for functional validation. The possibility of oligogenic inheritance suggests that combinatorial genetic effects may contribute to SCAD, requiring further investigation. Additionally, while pathways such as NOTCH/TGF crosstalk pathway and collagen biology emerged as potential contributors, experimental validation is necessary to confirm their mechanistic involvement. All cooccurring symptoms in the present study are self-reported, which should be taken into consideration when interpreting the data. Together, these limitations highlight the need for larger, multidisciplinary research efforts integrating genomics, functional studies, and deep phenotyping.

## Conclusion

This study supports SCAD as a genetically complex arteriopathy, driven by rare high-impact variants together with broader polygenic susceptibility. Variants in collagen, vascular extracellular matrix, and oestrogen-responsive pathways provide biologically plausible links to female-predominant disease. Although the diagnostic yield of clearly actionable variants is modest, these findings support broader genomic evaluation beyond overt syndromic presentations and highlight the need for larger integrative genomic and functional studies to refine risk stratification and management.

## Data Availability

The data will be available as suitable with Swedish law and the ethical approval

## Acknowledgements

We extend our sincere gratitude to all participating patients for their valuable contributions. We also thank Åsa Schippert and the Core Facility, a joint initiative between the Faculty of Medicine at Linköping University and Region Östergötland, for their assistance with DNA extraction from saliva samples and library preparation.

